# Did knowledge, attitudes, and practices matter during the second wave of COVID-19 pandemic in Bangladesh? Results from a web-based cross-sectional study

**DOI:** 10.1101/2023.05.02.23289398

**Authors:** Md. Robin Khan, Md. Jamal Hossain, Md. Ariful Islam, Shadid Uz Zaman, Mohammad Touhidul Islam, Md. Anamul Haque, Md. Rabiul Islam, Farhana Alam Ripa, Md. Monirul Islam, Foyez Ahmmed

## Abstract

Globally, health promotion measures have been undertaken in preventing the emergence and spread of coronavirus disease 2019 (COVID-19). However, whether these measures influence public awareness and behaviors is unclear and evidence is limited in particular in low-and-middle income country. We conducted an online survey among internet users in Bangladesh to understand the status and attributes of their knowledge, attitudes and practices towards COVID-19 during second wave of the pandemic when COVID educational information was more accessible to the public. Survey data were analyzed using descriptive statistics, Chi-square tests, and multivariate logistic regression analysis. Of 964 respondents, 40.2%, 51.5%, and 64.3% had good knowledge, confident attitudes, and proper practices towards COVID-19, respectively. The multivariate regression analysis found that the knowledge and practice scores were associated (p<0.05) with gender, age, and occupation. Females had better knowledge and practices compared to males (p<0.05). There were major gaps in awareness, attitudes, and practices among internet users in particular males and elders that needs to be addressed to control the further spread of COVID-19 infections before at least COVID-19 vaccine become accessible at population level in Bangladesh.

## Introduction

On 11^th^ March 2020, the World Health Organization (WHO) declared the novel coronavirus disease (COVID-19), caused by the pathogen severe acute respiratory syndrome coronavirus-2 (SARS-CoV-2), a global pandemic [1].This infection emerged as a zoonotic transmission from Wuhan, China, transmitted from animal to human and recognized as having a high ability to spread from human to human through direct touch and airway droplets. Symptoms include high fever, dry cough, breathing difficulties, and a loss of smell. And it may cause diarrhea, headaches, and body aches [2]. COVID-19 has spread to over 215 countries and territories around the world with more than 764 million confirmed cases and 6.9 million deaths [3].

Given that the treatment options are limited, public engagement and health education in outbreak responses have become important in managing outbreak responses [4]. Efforts include communicating what the disease entails and what symptoms to look out for, and changing people’s transmission-related behaviors to flatten the infection rates.

It is so important to emphasize that Bangladesh, being one of the most densely populated countries, carries a higher risk of spreading COVID-19.The first case of COVID-19 in the country was officially reported on 08 March 2020 and the first death was reported on18 March 2020 [5,6].The government has taken several preventative measures such as implementing lockdowns, limiting public transportation and endorsing quarantine, and spreading health awareness messages including maintenance of social distancing using massmedia [7]. Internet and social media are two important sources for people getting awareness messages towards preventing COVID-19 [8]. However, both the number of infected people and the death toll are rising; as of July 27, 2021, Bangladesh has officially recorded more than 1,179,827 cases and over 19,521 deaths [9].

Although several unprecedented strict measures were imposed by the government and international public health organizations since the inception of the ongoing pandemic, the success or collapse of these adopted attempts to impede the community transmission is directly dependent on the general people’s knowledge, attitudes, and practices (KAP) towards the disease. Besides, public behaviors and adherence to the preventive measures authorized by the authorities significantly influence the inhibition of the infection [10]. Furthermore, evidence endorses that public’ KAP is vital to battling public health emergencies such as COVID-19, and monkeypox these parameters are undeniably essential tools in identifying gaps and strengths of undertaken measures [10].

Researchers from multiple backgrounds providing exceptional efforts to explore the knowledge and practices in other regions of the world. For example, studies conducted in China, Africa, and Italy, identified trends where women, older and those of higher socioeconomic status were more knowledgeable, optimistic and had adopted appropriate practices towards COVID-19 [11–13].While in Saudi Arabia, the prevalence of KAP scores demonstrated a high level of knowledge, optimistic attitudes, and good level of practice towards the COVID-19 outbreak, while older adults and women had more knowledge about the disease [10]. Besides, a systematic review and meta-analysis revealed that the prevalence of average good knowledge, positive attitudes, and poor practice was 79.4%, 73.7%, and 40.3%, respectively, among the Ethiopian population during the COVID-19 outbreak that indicated a significant gap in KAP level in respect of WHO guided management and protective practices [14]. Misbeliefs and misconceptions about the SARS-CoV-2 infection and treatment persist among a significant percentage of people in Bangladesh [15].

Several studies conducted in Bangladesh have explored public’ KAP towards COVID-19 during the early phase with limited knowledge, poor attitudes, and practices tackling pandemic [16-23]. However, according to the best of our knowledge, very few studies have been conducted measuring people’s KAP towards COVID-19 during the second wave of COVID-19 in Bangladesh. A study conducted by Islam et al. [24] among the slum dwellers during the end of the first wave (August-September, 2020) of COVID-19 and reported that the majority of Bangladeshi slum dwellers had limited knowledge and poor practices (face mask and hand sanitization).

Maude et al. [25] investigated that a significant improvement in KAP level regarding the transmission of COVID-19 is possible by targetted promotional activities to address the gaps and limitations of KAP. By providing accurate information, it is possible to manage such crises and improve future pandemic preparedness. On the other hand, negative beliefs and behaviors toward newly infectious diseases can exacerbate epidemics, which may eventually lead to pandemics. The spread of the illness can be stopped when people work together to practice preventive behaviors including maintaining personal hygiene and avoiding close contact. For instance, altering behavior decreased the spread of the virus during the 2009 A/H1N1 influenza pandemic and the Zika outbreak [26]. Information raises citizens’ knowledge levels and promotes positive attitudes, which may aid in the fight against COVID-19 and other potential threats like monkeypox in the future. Since, SARS-CoV-2 is a novel virus with a high mutation power, which has resulted in multiple variants and waves of COVID-19. New variants with new waves won’t come as a surprise to us because the world is still fighting the disease with new cases emerging every day. In Bangladesh, where daily cases of covid 19 is still present, it is imperative to investigate how the public has been responding to the awareness messages and recommended practices to avoid developing COVID-19 and prepared themselves to fight with the other infectious disease like monkeypox. This study explored the knowledge, attitudes, and practices (KAP) of active internet users in Bangladesh towards COVID-19 during the second wave of the pandemic to identify relevant gaps and inform health promotion strategies, including target groups to effectively control COVID-19 in Bangladesh in years ahead.

## Methods

### Study design

A multi-variable, cross-sectional survey has been conducted amongst active internet users in Bangladesh. “Active” internet users were defined as any individual who had an internet footprint, i.e. a social media handle or an active email address; these internet accounts were later used to confirm if the participant was legitimate. The questionnaire was prepared using Google Forms and the link was shared in the contact lists of the investigators and to WhatsApp and Facebook groups. The respondents were also encouraged to disseminate survey link with their peers, a snowball sampling technique [27-29] was utilized to recruit participants. A resident of Bangladesh with access to the internet, and a willingness to engage were requirements for inclusion in the study. People who did not actively consent to the survey and who did not provide their social media account, phone number, or email address for data verification were omitted. Other exclusion criteria included being under the age of 18. Responses were collected between December 28, 2020 and January 3, 2021.

### Survey questionnaire

The survey questionnaire consisted of four sections. The first section was the scope of the study and ethical concerns. The second section collected personal and demographic data. The third and fourth sections of the questionnaire focused on the knowledge, attitudes, and practices (KAP) of the respondents regarding COVID-19. The questionnaire used for data collection was based on the report published by Zhong et al. [11]. A total of thirteen questions were related to knowledge and five questions covered attitudes and practices. Among the comprehensive knowledge-based questions, two questions bear general information of the COVID-19 outbreak, three were transmission-related questions, two questions were symptoms-related, four questions were preventive measures associated questions, and finally, the rest of the two questions were treatment and physical complications related. To validate the questionnaire, it was implemented in two steps. Firstly, we disseminated our initial draft to the team investigators and professional colleagues for review on the significance and comprehensiveness of the survey. Using the feedback, we added Bengali (Bangla) translations and made them more simplistic for the general public. The translation was validated through a forward and backward translation method with the help of a bilingual expert and who have enough knowledge about medical terminology [29]. Thereafter, 53 persons took part in the questionnaire’s pilot testing before a final version was made in order to evaluate its reliability. Data from the pilot study were imported into IBM SPSS Version 20 and reliability coefficient analysis was done. According to the pilot data, the knowledge, attitude, and practice showed respective Cronbach’s alpha coefficients of 0.73, 0.68, and 0.71, which shows the internal reliability of the questionnaire.The data from this pilot study was not used in our final results.

### Variables and Scoring

Several demographic parameters were used to analyze the knowledge, attitudes, and practices of the population under study. These parameters included gender, age, marital status, residence, occupation, and primary social media. The responses for the KAP questions were scored separately for each variable. The correct answer to a question was scored as “1”, an incorrect and not sure answer was scored as “0” for knowledge score, whereas the options were ‘agree’ = ‘1’, ‘disagree’ = ‘0’ and ‘not sure’ = ‘0’ for attitudes-based questions [23, 30]. Besides, the options were ‘yes’ and ‘no’ for the practice-related questions. Using this scoring methodology, the knowledge score, attitude score, and practice score were determined for the categories under each variable. The scoring system was adopted based on the available knowledge uncovered thus far and disseminated by reliable authorities like the World Health Organization (WHO), the center for disease control (CDC) of the United States, and the National Health Service (NHS) of England. The knowledge was categorized as “good” or “poor” based on the score; similarly, the attitude was categorized as “confident” or “confused,” while practices were categorized as either “proper” or “improper”. Participants who correctly answered at least 10 questions out of 13 (nearly 80%, according to Bloom’s cut-off score [23, 31]) were considered to have “Good” knowledge regarding COVID-19, otherwise considered to have “Poor” knowledge. Besides, participants who showed positive responses to both attitude-related questions were considered to be “Confident”; otherwise, they were considered to be “Confused”. Furthermore, participants who provided positive responses to all three practice-related questions were maintaining “Proper” practice; otherwise, they were considered to maintain “Improper” practice.

### Statistical Analysis

A chi-square (χ^2^) test was performed to evaluate the association between the knowledge, attitude, and practice with different background characteristics of the study participants. Multivariate logistic regression analysis was conducted to find the potential socioeconomic and demographic factors associated with knowledge of COVID-19 and the practice of sanitation. The reference categories for the regression analysis were indicated by “R”. Results were illustrated as an adjusted odd ratio (AOR) and 95% confidence interval (CI) with a significance level (*p*< 0.05) [32]. In addition, *t*-test and one-way analysis-of-variance (ANOVA) were used to determine any potential differences in mean scores of KAP among the categories within a variable. All analyses were performed using IBM SPSS Version 20.

### Participants, ethics, and approval

The Bangladeshi active internet users ≥ 18 years of age having social media ID or Email address, and who clearly understood the questionnaire were eligible to respond to the survey. In the introduction section of the questionnaire, the participants were clearly introduced to the study’s aim and objective. Besides, all the respondents were ensured that their provided information and opinions would have remained confidential and anonymous. Furthermore, the survey included a consent section where the participants confirmed their virtual consent and willingness for voluntary participation with no reward and payment. Finally, the research’s procedure and detailed protocols were critically reviewed and approved by the ethics committee (Ref. No. DoP/RC/EC/2020/06/app/01OL), the Department of Pharmacy, the Northern University of Bangladesh under the condition of complete adherence to the Declaration of Helsinki [33].

## Results

### Data

A total of 1396 responses were collected during the study period. Among these responses, 1286 responses were verified to be unquestionably authentic because some respondents failed to answer all questions or provide their social media account, phone number, or e-mail address. Then, 75% of the data i.e. 964 responses were selected randomly using a randomization protocol. All analyses described below were performed on these 964 responses.

### Socioeconomic and Demographic Distribution of the participants

Univariate analysis was performed on the data (N = 964) to understand the socioeconomic and demographic distribution of the participants. The socioeconomic and demographic parameters considered were gender, age, region of residence, marital status, occupation, the effect of COVID-19 on the income status, and the primary source of information on the Internet. Among the participants, 97.2% were between the ages of 18 and 37 years, 51.56% were male and 48.44% were female, and most of them were unmarried (86.10%). The majorities within the other categories consisted of 71.27% residing in the urban parts of Bangladesh and 76.76% were students. Lastly, the income status did decrease for 77.18% of the sample size and 79.25% of our respondents use Facebook as their primary social media account. The summary of the data has been shown in **Table 1**.

**Table 1:**
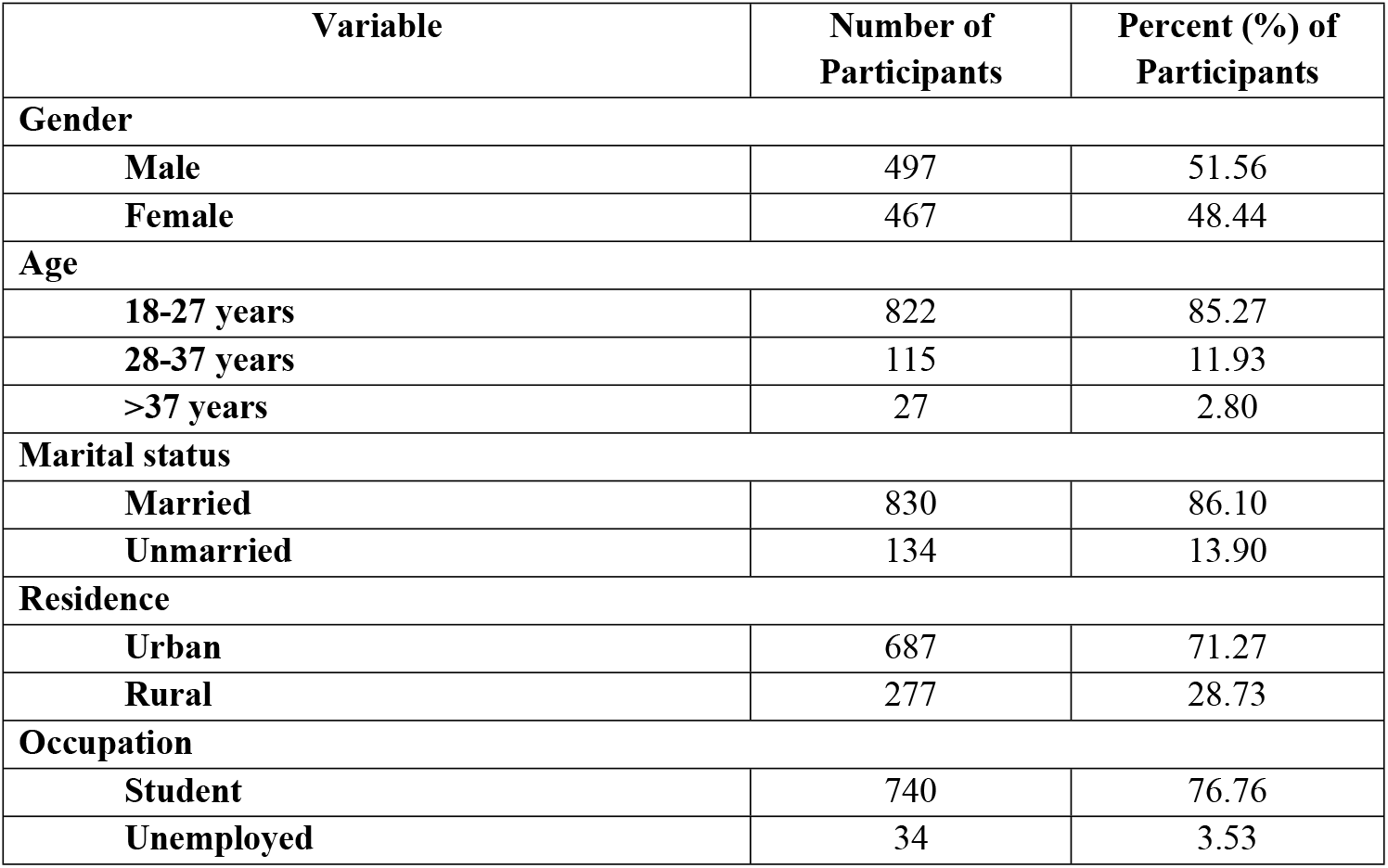
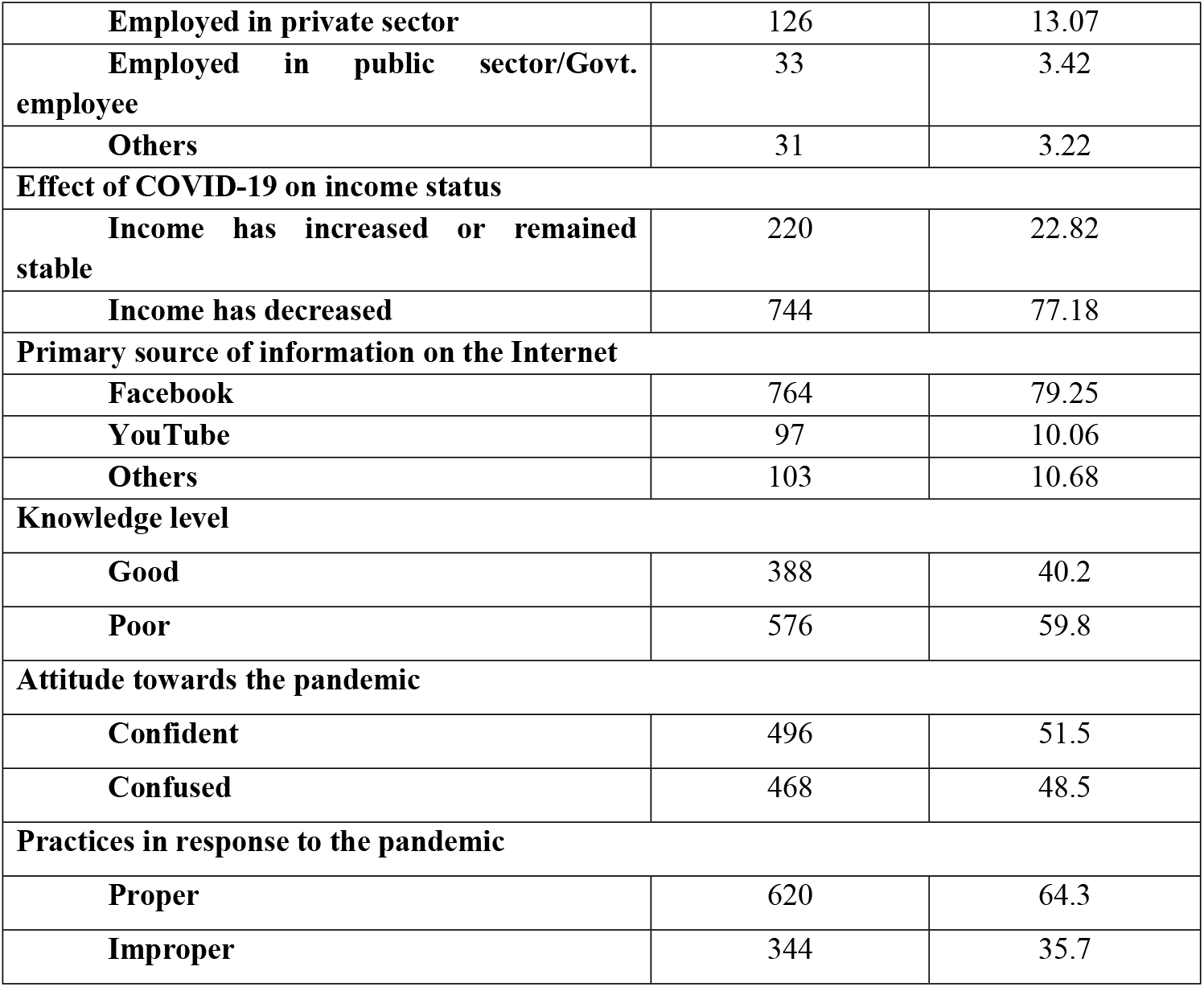
Sociodemographic and KAP distributions of the active internet users during the second wave of COVID-19 outbreak in Bangladesh.

### Knowledge, attitides, and practice (KAP)

The goal of this research was to examine the level of knowledge possessed by the participants regarding COVID-19, their attitude towards the pandemic, and their practices in response to the pandemic. Fig 1 illustratedthe distribution of the participants under these categories. From the univariate analysis, the prevalence of good knowledge, confident attitudes, and proper practice were 40.2% (n = 388), 51.5% (n = 496), and 64.3% (n = 620), respectively among the active internet users during the second wave of COVID-19 Bangladesh. In contrast, a significant percentage of the participants obtained poor knowledge scores (59.8%) and showed confused attitudes (48.5%) and improper practices (35.7%) (Table 1).

**Fig 1.**
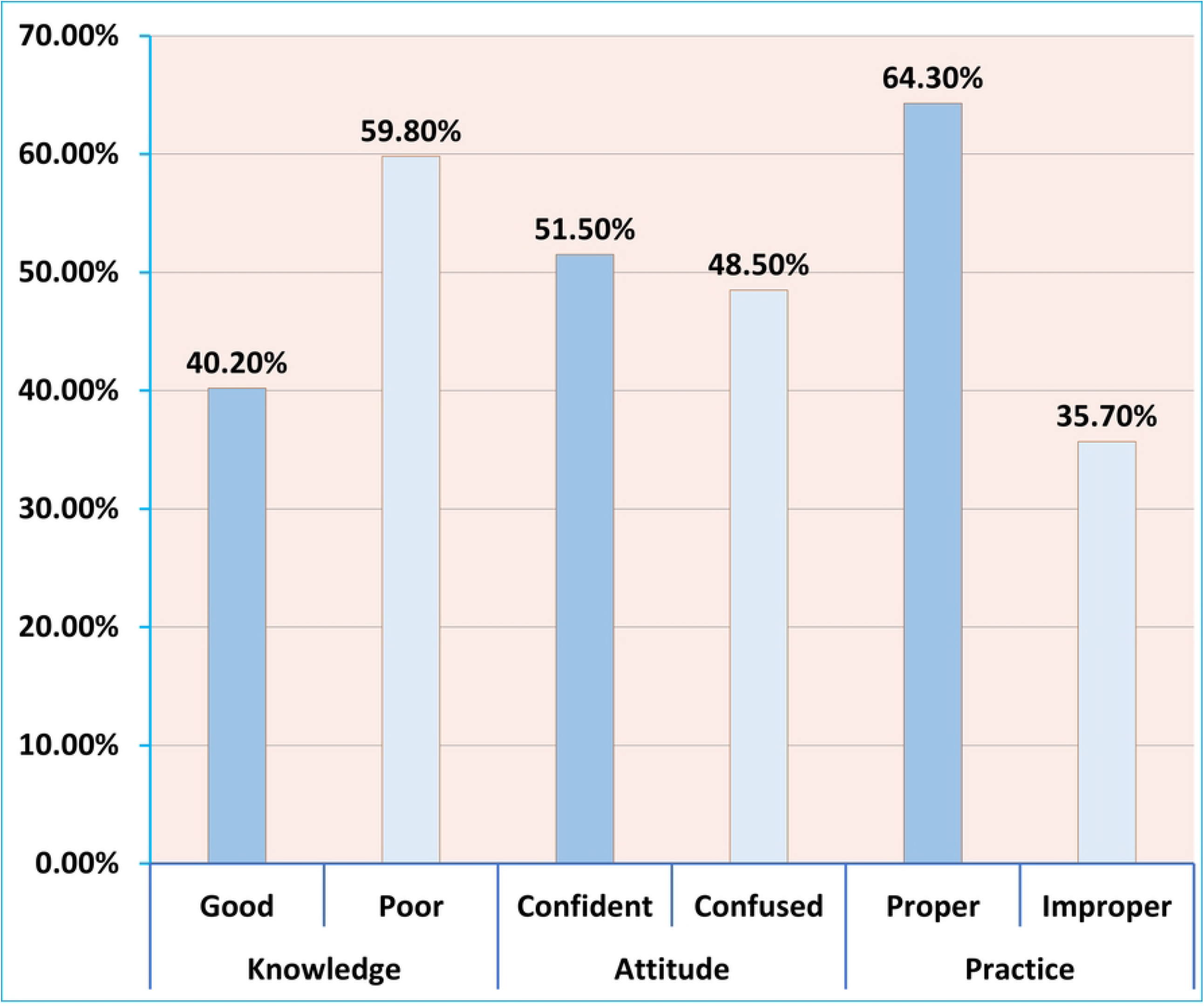
**Percentage distributions of good and poor knowledge, confident and confused attitudes, and proper and improper practice among the internet users during the second wave of COVID-19 outbreak in Bangladesh.**

### Chi-sqaure (χ^2^) analysis

Bivariate analysis, specifically a χ^2^ test of independence, was performed to determine any potential association between the socioeconomic or demographic variables and the knowledge, attitude, and practices scores. The findings obtained from this bivariate analysis were summarized in Table 2.

**Table 2:**
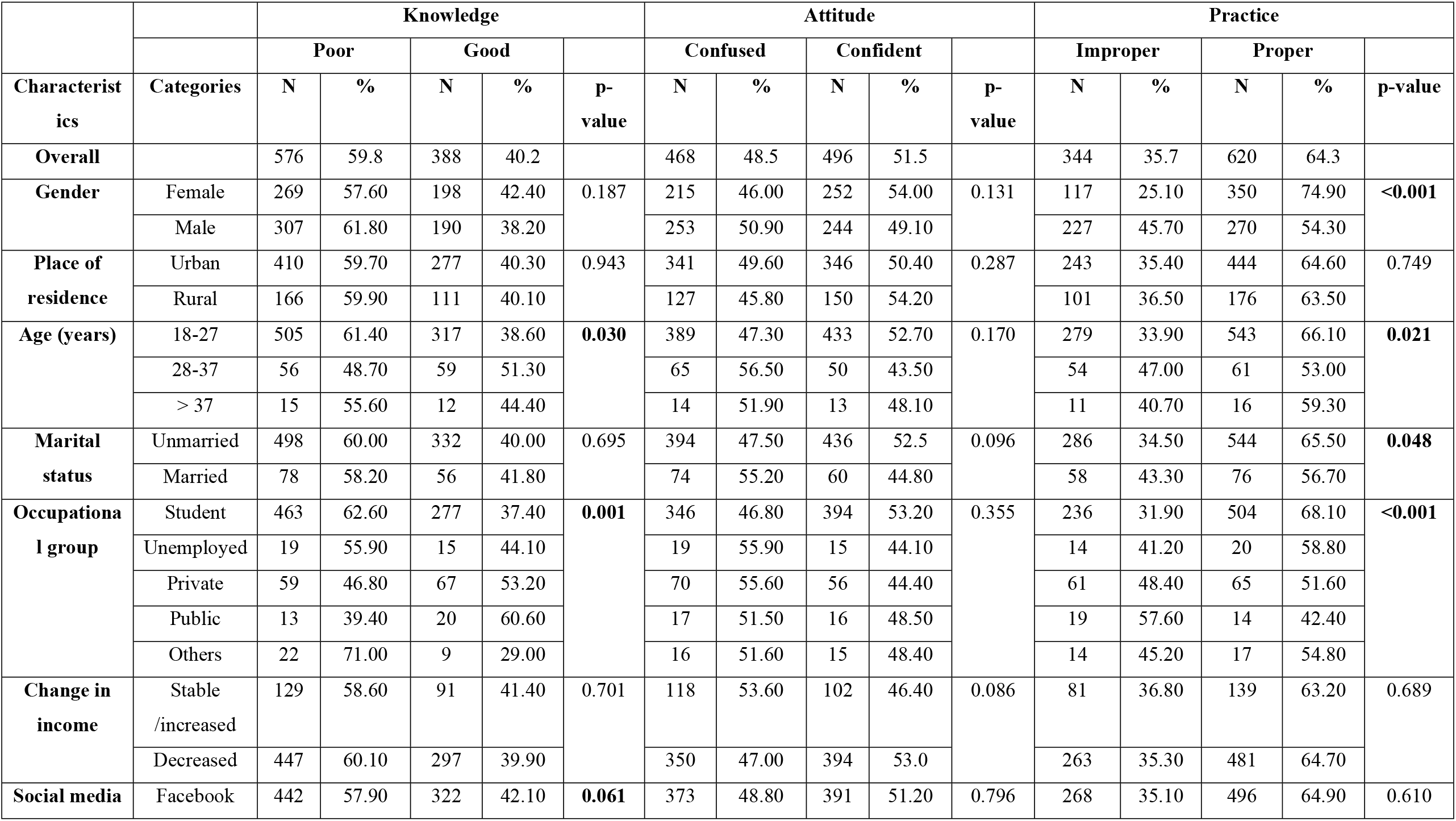
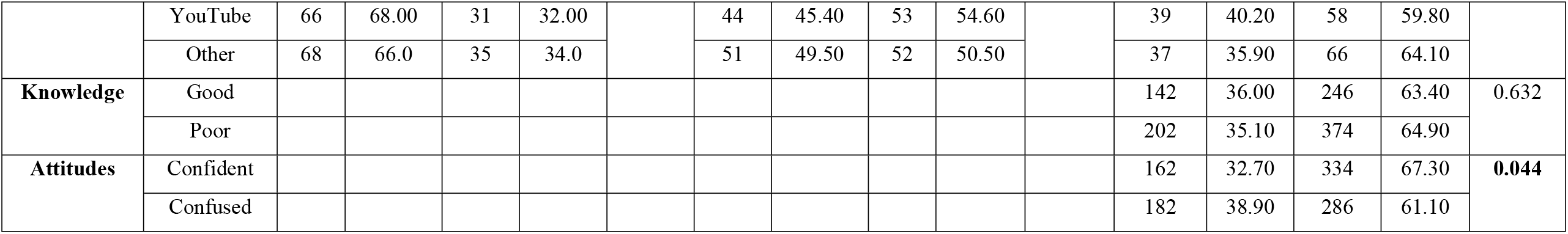
Chi-square (χ^2^) test for finding theassociation of socioeconomic and demographic characteristics of respondents with knowledge, attitude, and practice among the active users during the second wave of COVID-19 outbreak in Bangladesh.

|  |  | Knowledge |  |  |  |  | Attitude |  |  |  |  | Practice |  |  |  |  |
| --- | --- | --- | --- | --- | --- | --- | --- | --- | --- | --- | --- | --- | --- | --- | --- | --- |
|  |  | Poor |  | Good |  | p-value | Confused |  | Confident |  | p-value | Improper |  | Proper |  | p-value |
| Characteristics | Categories | N | % | N | % |  | N | % | N | % |  | N | % | N | % |  |
| Overall |  | 576 | 59.8 | 388 | 40.2 |  | 468 | 48.5 | 496 | 51.5 |  | 344 | 35.7 | 620 | 64.3 |  |
| Gender | Female | 269 | 57.60 | 198 | 42.40 | 0.187 | 215 | 46.00 | 252 | 54.00 | 0.131 | 117 | 25.10 | 350 | 74.90 | <0.001 |
|  | Male | 307 | 61.80 | 190 | 38.20 |  | 253 | 50.90 | 244 | 49.10 |  | 227 | 45.70 | 270 | 54.30 |  |
| Place of residence | Urban | 410 | 59.70 | 277 | 40.30 | 0.943 | 341 | 49.60 | 346 | 50.40 | 0.287 | 243 | 35.40 | 444 | 64.60 | 0.749 |
|  | Rural | 166 | 59.90 | 111 | 40.10 |  | 127 | 45.80 | 150 | 54.20 |  | 101 | 36.50 | 176 | 63.50 |  |
| Age (years) | 18-27 | 505 | 61.40 | 317 | 38.60 | 0.030 | 389 | 47.30 | 433 | 52.70 | 0.170 | 279 | 33.90 | 543 | 66.10 | 0.021 |
|  | 28-37 | 56 | 48.70 | 59 | 51.30 |  | 65 | 56.50 | 50 | 43.50 |  | 54 | 47.00 | 61 | 53.00 |  |
|  | > 37 | 15 | 55.60 | 12 | 44.40 |  | 14 | 51.90 | 13 | 48.10 |  | 11 | 40.70 | 16 | 59.30 |  |
| Marital status | Unmarried | 498 | 60.00 | 332 | 40.00 | 0.695 | 394 | 47.50 | 436 | 52.5 | 0.096 | 286 | 34.50 | 544 | 65.50 | 0.048 |
|  | Married | 78 | 58.20 | 56 | 41.80 |  | 74 | 55.20 | 60 | 44.80 |  | 58 | 43.30 | 76 | 56.70 |  |
| Occupational group | Student | 463 | 62.60 | 277 | 37.40 | 0.001 | 346 | 46.80 | 394 | 53.20 | 0.355 | 236 | 31.90 | 504 | 68.10 | <0.001 |
|  | Unemployed | 19 | 55.90 | 15 | 44.10 |  | 19 | 55.90 | 15 | 44.10 |  | 14 | 41.20 | 20 | 58.80 |  |
|  | Private | 59 | 46.80 | 67 | 53.20 |  | 70 | 55.60 | 56 | 44.40 |  | 61 | 48.40 | 65 | 51.60 |  |
|  | Public | 13 | 39.40 | 20 | 60.60 |  | 17 | 51.50 | 16 | 48.50 |  | 19 | 57.60 | 14 | 42.40 |  |
|  | Others | 22 | 71.00 | 9 | 29.00 |  | 16 | 51.60 | 15 | 48.40 |  | 14 | 45.20 | 17 | 54.80 |  |
| Change in income | Stable /increased | 129 | 58.60 | 91 | 41.40 | 0.701 | 118 | 53.60 | 102 | 46.40 | 0.086 | 81 | 36.80 | 139 | 63.20 | 0.689 |
|  | Decreased | 447 | 60.10 | 297 | 39.90 |  | 350 | 47.00 | 394 | 53.0 |  | 263 | 35.30 | 481 | 64.70 |  |
| Social media | Facebook | 442 | 57.90 | 322 | 42.10 | 0.061 | 373 | 48.80 | 391 | 51.20 | 0.796 | 268 | 35.10 | 496 | 64.90 | 0.610 |
|  | YouTube | 66 | 68.00 | 31 | 32.00 |  | 44 | 45.40 | 53 | 54.60 |  | 39 | 40.20 | 58 | 59.80 |  |
|  | Other | 68 | 66.0 | 35 | 34.0 |  | 51 | 49.50 | 52 | 50.50 |  | 37 | 35.90 | 66 | 64.10 |  |
| Knowledge | Good |  |  |  |  |  |  |  |  |  |  | 142 | 36.00 | 246 | 63.40 | 0.632 |
|  | Poor |  |  |  |  |  |  |  |  |  |  | 202 | 35.10 | 374 | 64.90 |  |
| Attitudes | Confident |  |  |  |  |  |  |  |  |  |  | 162 | 32.70 | 334 | 67.30 | 0.044 |
|  | Confused |  |  |  |  |  |  |  |  |  |  | 182 | 38.90 | 286 | 61.10 |  |

The age group and occupational status were significantly associated with knowledge level (Table 2). Respondents from 28-37 years age group exhibited the highest percentage in having good knowledge (51.30%) compared to 18-27 years (38.60%) and those older than 37 years (44.40%) groups. Besides, public job holders dominated in having good knowledge (60.60%) followed by private (53.20%), unemployed (44.10%), students (37.40%), and others (29%).

In the case of attitude, there was no significant association at 5% confidence interval but marital status (p=0.096) and change in income (p=0.086) showed significant association at 10% confidence interval. Positive attitudes were the highest amongst those who were female (54.0%), lived in rural areas (54.20%), and students (53.20%), as well as those who were unmarried (52.5%) and between the ages of 18-27 (52.70%).

We assessed the practice of public such as avoiding crowded areas, wearing facial masks when leaving their homes, and practicing proper sanitation methods. In our study, we found females practiced proper methods (74.90%) compared to males (54.30%), and gender was found significantly (p<0.01) associated with the practice. We also found that age groups, marital status, and occupational groups were significantly associated (p<0.05) with the practice of sanitation. Respondents aged 18-27 years (66.10%) practiced sanitations more properly followed by more than 37 (59.30%) and 28-37 (53.00%) age groups. Besides, the students (68.10%) followed proper practices, followed by the unemployed (58.80%), others (54.80%), private (51.60%), and the public (42.40%) job holders. Although it was expected that those who had increasingly good knowledge were going to exhibit proper hygiene methods, we did not get a significant association between knowledge and practice. Interestingly, we were able to see a significant association between attitude and practice— respondents who were confident about the recovery from the pandemic had a higher percentage of proper practice (67.30%) compared to their opposite counterparts. Similarly, all the findings from χ^2^ analysis were tabulated in Table 2.

### Logistic regression analysis

A logistic regression analysis was employed during multivariate analysis to confirm the potential factors associated with good knowledge and proper practice after adjusting other factors, and the outcomes obtained from the analysis were tabulated in Table 3. It is noted here that the trends in attitude were observed to be about half the subcategory demonstrating positive attitudes towards the effectiveness of Bangladesh’s government control of the coronavirus. We did not consider regression analysis for attitude as we did not get a significant association for any of the covariates from the chi-square test.

**Table 3:**
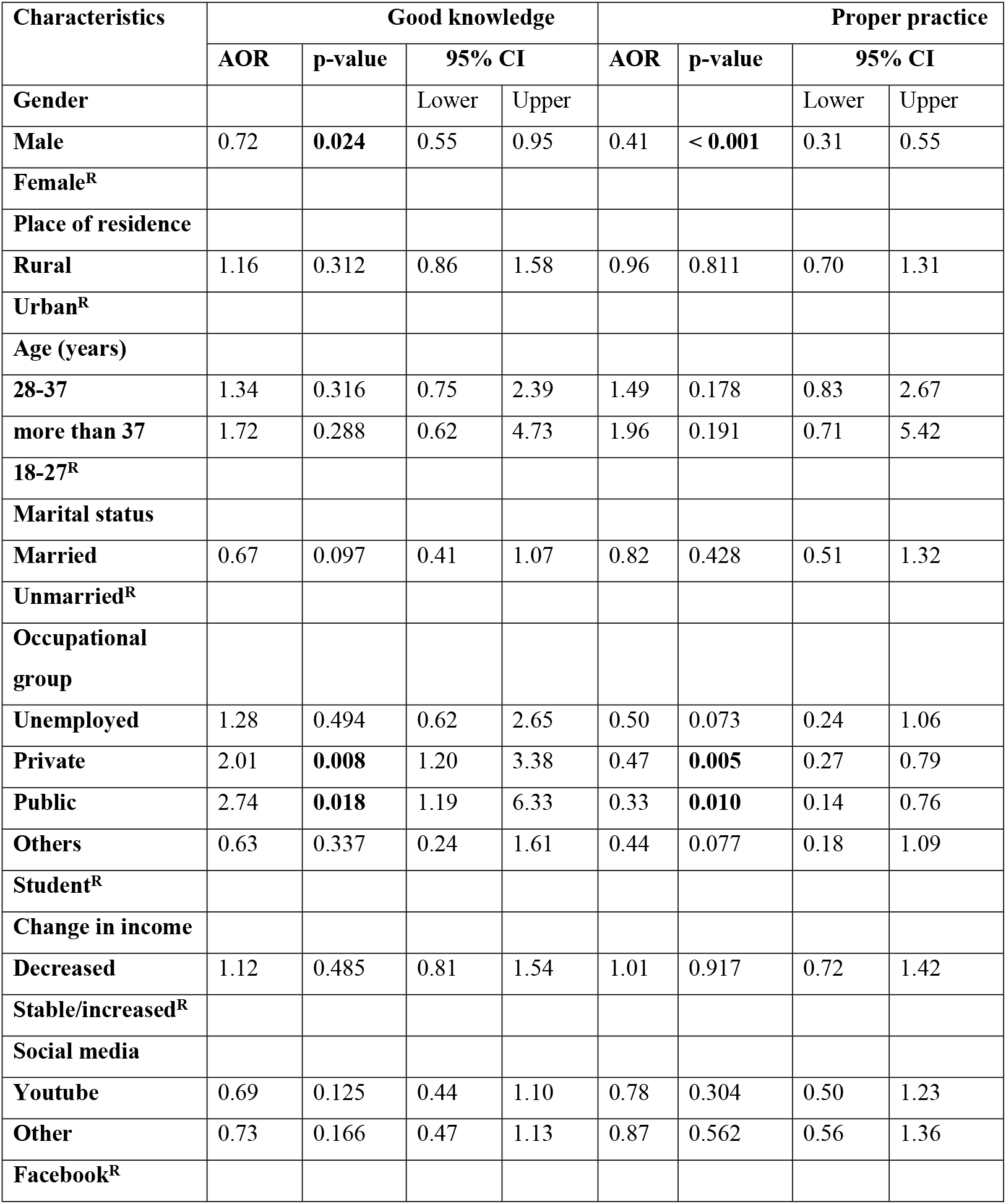
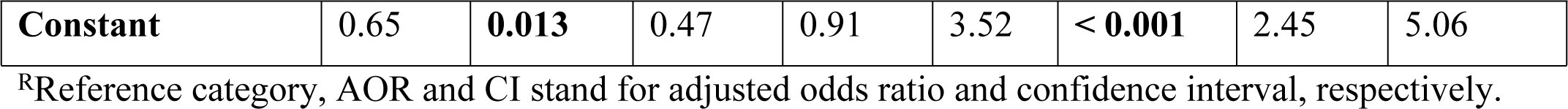
Multivariate logistic regression analysis for finding the potential socioeconomic and demographic factors associated with good knowledge and proper practice among the populations during the second wave of COVID-19 outbreak in Bangladesh.

| Characteristics | Good knowledge |  |  |  | Proper practice |  |  |  |
| --- | --- | --- | --- | --- | --- | --- | --- | --- |
|  | AOR | p-value | 95% CI |  | AOR | p-value | 95% CI |  |
| Gender |  |  | Lower | Upper |  |  | Lower | Upper |
| Male | 0.72 | **0.024** | 0.55 | 0.95 | 0.41 | **< 0.001** | 0.31 | 0.55 |
| Female<sup>R</sup> |  |  |  |  |  |  |  |  |
| Place of residence |  |  |  |  |  |  |  |  |
| Rural | 1.16 | 0.312 | 0.86 | 1.58 | 0.96 | 0.811 | 0.70 | 1.31 |
| Urban<sup>R</sup> |  |  |  |  |  |  |  |  |
| Age (years) |  |  |  |  |  |  |  |  |
| 28-37 | 1.34 | 0.316 | 0.75 | 2.39 | 1.49 | 0.178 | 0.83 | 2.67 |
| more than 37 | 1.72 | 0.288 | 0.62 | 4.73 | 1.96 | 0.191 | 0.71 | 5.42 |
| 18-27<sup>R</sup> |  |  |  |  |  |  |  |  |
| Marital status |  |  |  |  |  |  |  |  |
| Married | 0.67 | 0.097 | 0.41 | 1.07 | 0.82 | 0.428 | 0.51 | 1.32 |
| Unmarried<sup>R</sup> |  |  |  |  |  |  |  |  |
| Occupational group |  |  |  |  |  |  |  |  |
| Unemployed | 1.28 | 0.494 | 0.62 | 2.65 | 0.50 | 0.073 | 0.24 | 1.06 |
| Private | 2.01 | **0.008** | 1.20 | 3.38 | 0.47 | **0.005** | 0.27 | 0.79 |
| Public | 2.74 | **0.018** | 1.19 | 6.33 | 0.33 | **0.010** | 0.14 | 0.76 |
| Others | 0.63 | 0.337 | 0.24 | 1.61 | 0.44 | 0.077 | 0.18 | 1.09 |
| Student<sup>R</sup> |  |  |  |  |  |  |  |  |
| Change in income |  |  |  |  |  |  |  |  |
| Decreased | 1.12 | 0.485 | 0.81 | 1.54 | 1.01 | 0.917 | 0.72 | 1.42 |
| Stable/increased<sup>R</sup> |  |  |  |  |  |  |  |  |
| Social media |  |  |  |  |  |  |  |  |
| Youtube | 0.69 | 0.125 | 0.44 | 1.10 | 0.78 | 0.304 | 0.50 | 1.23 |
| Other | 0.73 | 0.166 | 0.47 | 1.13 | 0.87 | 0.562 | 0.56 | 1.36 |
| Facebook<sup>R</sup> |  |  |  |  |  |  |  |  |
| **Constant** | 0.65 | **0.013** | 0.47 | 0.91 | 3.52 | **< 0.001** | 2.45 | 5.06 |

<sup>R</sup>Reference category, AOR and CI stand for adjusted odds ratio and confidence interval, respectively.

From the multivariate regression analysis, we found that male respondents had 28% of lower odds of having good knowledge than the female respondents (adjusted odds ratio [AOR] = 0.72, 95% CI = 0.55 to 0.95; p < 0.024). We also observed that private job holders (AOR= 2.01, p <0.01) and public job holders (AOR = 2.74, p <0.05) were more likely to have good knowledge than the students.

In case of practice, the female participants exerted more proper practice compared to the male particpants in Bangladesh (male vs. female: AOR = 0.41, 95% CI = 0.31 to 0.55; p<0.001). Besides, the students exhibited more proper practice than both private (vs. students: AOR = 0.47, 95% CI = 0.27 to 0.79; p = 0.005) and public (vs. students: AOR = 0.33, 95% CI = 0.14 to 0.76; p < 0.01) sector employees. Similarly, all the results extrapolated from the regression analysis were summazired in Table 3.

### Measurement of Misconception

The data collected from the participants were also used to make a quantitative assessment of misconceptions among the participants. For this purpose, the mean number of wrong answers given in the knowledge segment of the questionnaire was determined for each variable. Consequently, *t*-test or one-way ANOVA was performed to determine whether any statistically significant differences existed between categories within each variable. It was observed that male participants provided more wrong answers in the knowledge segment compared to female participants (p<0.05). Statistically significant difference in the mean number of wrong answers were also observed among various occupational groups with participants employed in the public sector providing least number of wrong answers compared to other groups such as students, private employees, and the unemployed (**Table 4)**.

**Table 4:**
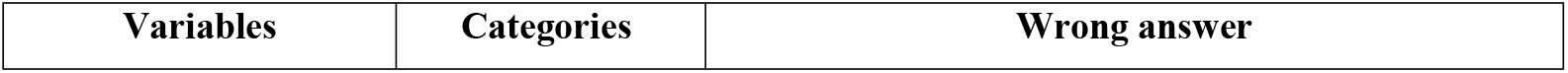
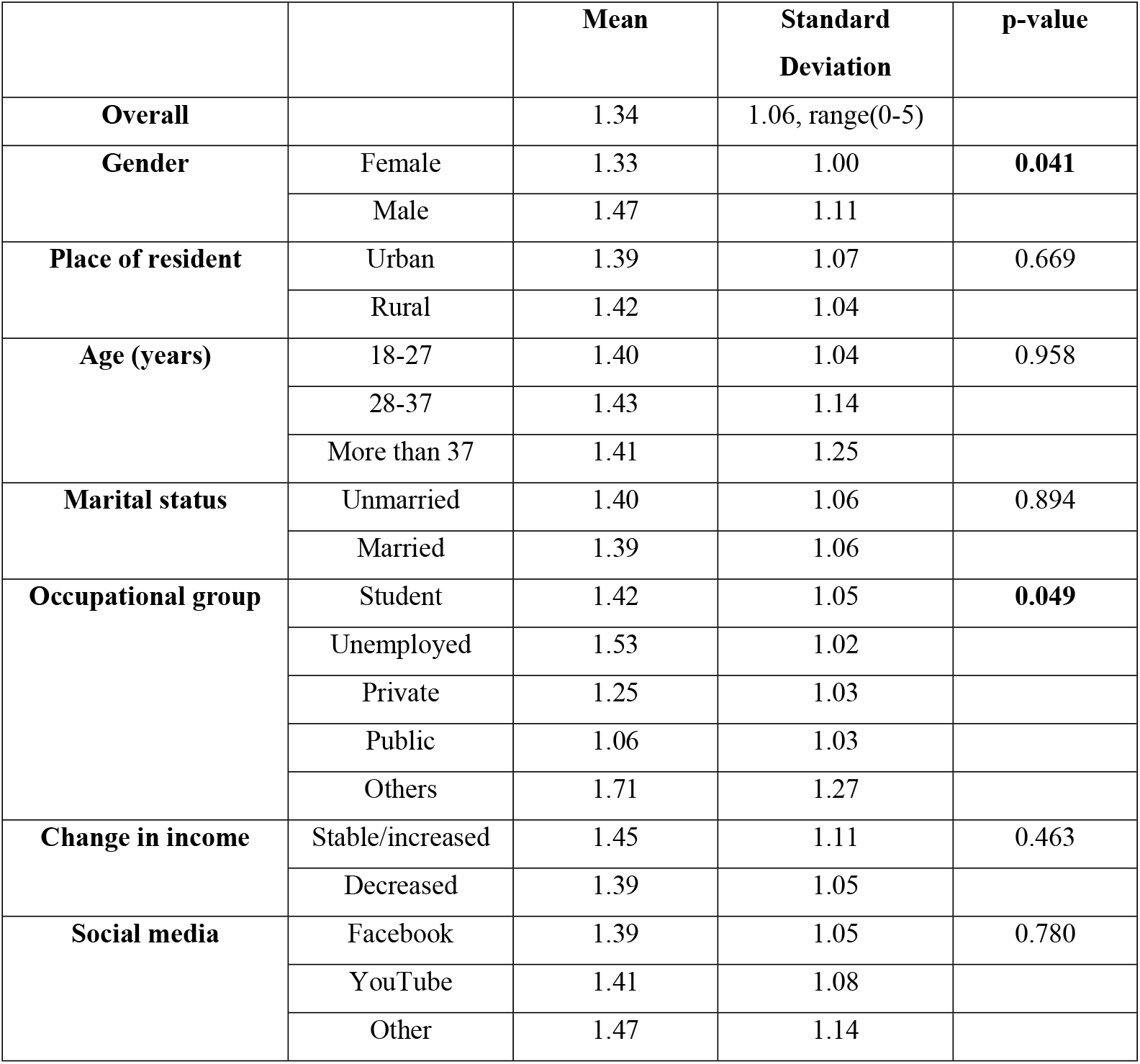
Difference in average number of wrong answers by different background characteristics of respondents. p-values obtained from independent sample t-test and one-way ANOVA.

|  |  | Mean | Standard Deviation | p-value |
| --- | --- | --- | --- | --- |
| **Overall** |  | 1.34 | 1.06, range(0-5) |  |
| **Gender** | Female | 1.33 | 1.00 | **0.041** |
|  | Male | 1.47 | 1.11 |  |
| **Place of resident** | Urban | 1.39 | 1.07 | 0.669 |
|  | Rural | 1.42 | 1.04 |  |
| **Age (years)** | 18-27 | 1.40 | 1.04 | 0.958 |
|  | 28-37 | 1.43 | 1.14 |  |
|  | More than 37 | 1.41 | 1.25 |  |
| **Marital status** | Unmarried | 1.40 | 1.06 | 0.894 |
|  | Married | 1.39 | 1.06 |  |
| **Occupational group** | Student | 1.42 | 1.05 | **0.049** |
|  | Unemployed | 1.53 | 1.02 |  |
|  | Private | 1.25 | 1.03 |  |
|  | Public | 1.06 | 1.03 |  |
|  | Others | 1.71 | 1.27 |  |
| **Change in income** | Stable/increased | 1.45 | 1.11 | 0.463 |
|  | Decreased | 1.39 | 1.05 |  |
| **Social media** | Facebook | 1.39 | 1.05 | 0.780 |
|  | YouTube | 1.41 | 1.08 |  |
|  | Other | 1.47 | 1.14 |  |

## Discussion

During the second wave of COVID 19, the study aimed to evaluate the knowledge, attitudes, and practice of Bangladeshi internet users. The results showed a significant number of sociodemographic characteristics that influence knowledge, attitude, and practice and should be helpful when developing health education programs regarding newly emerging infectious diseases. 40.2% of the group polled showed good knowledge, 51.5% showed confident attitudes, and 64.3% adhere to effective COVID-19 practices. On the other hand, there was significant correlation between knowledge, practice with gender, age, and occupation groups.Bangladesh Telecommunication Regulatory Commission does not publish any demographic data on the internet users, and data from other sources regarding this is scarce. However, Islam et al. [8] have recently reported that around 45 million (27.2%) people out of 168 million are currently using various social media platform in Bangladesh [34].Notably, in neighboring country India, 71% of the total internet users were between the age of 16 and 39, while less than 50% of the users lived in urban areas as of November, 2019 [35]. It is difficult to make any comparison to the data obtained in this study with that of the data from India, but it is mentionable that the data obtained in this study is more urban-centric. For 80% of the participants, Facebook is the primary source of information on the internet. This may be potentially alarming because a research previously pointed out that misleading information regarding Zika virus was more popular than useful information on Facebook and had greater dissemination [36]. Another research of the 492 respondents revealed that social media (107/492; 21.8%) and television (264/492, 53.7%) are the two main sources of information for Bangladeshi people regarding the COVID 19 [37]. Social media users of Bangladesh are not only cofined into Facebook they also use YouTube, Twitter, WhatsApp, LinkedIn, Instagram, Reddit, and Tumblr, among other sites. An online survey that included 265 (N=265) Bangladeshi respondents of various ages from June to October 2020 revealed that creative social media use improves knowledge of Covid-19 precautions, and that this important knowledge aids in preventing Covid-19 outbreak in Bangladesh [38].

According to the scoring methodology adopted in this study, the enumerated good knowledge, confident attitudes, and proper practice level among the active internet users were on par with or higher than the reported by several studies conducted in Bangladesh [16-23]. The good knowledge levels were around 48% and 61%, according to Ferdous et al. [21] and Banik et al. [23], respectively, for Bangladeshi populations. Besides, the study conducted by Hossain et al. [17] revealed the level of positive attitudes was 48.25% that might be equivalent to the current report. In terms of practices, 97.20% followed recommended hygiene practices like washing hands and avoiding unnecessary touches, while 66.80% avoided crowded places (data not shown in the Results section). The percentage of wearing the mask during the early phase COVID-19 in Bangladesh was below 70% [17]. In contrast, the study found that 98.34% of the participants wore masks when leaving the household, indicating a clear improvement in proper practices among Bangladesh populations during the second wave of COVID-19 outbreak. The differences between this study and the other studies conducted in Bangladesh may be attributed to the fact that the other studies were conducted during March and April of 2020, when infection had just started spreading in the country and there was less information available on the disease. There are also significant differences in the questionnaire design and scoring methodology, which may also be a factor.

Moreover, the percentage of participants with good knowledge and good practices obtained in this study are lower than that obtained from a study conducted in Pakistan[39]. A similar study conducted in India showed that there was a considerable misconception among the participants; however, the study did not categorize its participants as having good knowledge or poor knowledge [40]. Studies conducted in other Asian countries like Malaysia, Saudi Arabia, and Iran reported a higher percentage of participants with good knowledge compared to this study [10,41,42]. Similar observations can be made regarding the percentage of participants with good practices. However, due to differences in reporting, a direct comparison is difficult to make. Furthermore, these studies collected data in the early phase of COVID-19 pandemic, during February and March. The percentage of participants with good knowledge in this study is also lower than those obtained from Egypt, Nigeria, Sudan, and Ethiopia; however, comparatively lower percentage of participants maintained good practices (e.g. wearing masks) in these studies [12,43–45].The Lower percentage of people with good practices in these studies compared to the current study may be attributed to economic conditions in the respective countries among other things.

An important aspect of this study was the evaluation of the association between knowledge, attitude, and practices. Bivariate analysis of data via χ^2^test revealed that confident attitude towards COVID-19 was significantly associated with good practices however no statistically significant association was found between knowledge and practices or knowledge and attitude. However, Ferdous et al. [21] described a statistically significant association between attitude and practices, while Paul et al. [16]actually found that many participants with good knowledge had poor attitudes and practices. Conversely, in the studies conducted in China, higher knowledge level was found to be associated with positive attitude,[11] though one study described poor practices and poor attitude in the rural participants compared to urban participants[46].In Middle Eastern countries like Saudi Arabia, knowledge was found to have statistically significant association with both practices and attitudes,[10] and among Irani participants, there was 37% correlation between knowledge and practice [42]. Studies conducted in Egypt and Nigeria showed that knowledge was significantly associated with attitude/perception,[12, 44] while study on the Sudanese population showed that higher knowledge had statistically significant association with both positive attitude and good practices [43].

Multivariate analysis also found that male participants are statistically less knowledgeable than females. In addition, it was also observed that males provided significantly more wrong answers than females in this study. A similar result was found in the study by Wadoodet al.[47]. In contrast, Ferdous et al. [21] and Paul et al. [16] found no statistically significant gender difference in knowledge while Hossainet al. [19] 2020 actually found that males were more knowledgeable than females.Outside Bangladesh, studies conducted by Al-Hanawiet al. [10] (Saudi Arabia), Abdelhafizet al. [48] (Egypt), Aynalemet al. [49] (Ethiopia, students)found no statistically significant gender difference in knowledge level. Besides, Azlanet al. [41] (Malaysia), Hager et al. [12] (Nigeria and Egypt), Hezimaet al. [43] (Sudan), Honarvaret al. [42] (Iran), and so on, reported that either the knowledge level of the female respondents was higher than males or unsatisfactory knowledge level was more pronounced in males than in females. Only Hayat et al. [39] 2020 reported males having higher knowledge level compared to female respondents in Pakistan. Consequently, it has appeared that not only in Bangladesh, but also in many other parts of the world, females were more knowledgeable about COVID-19 than males.

Paradoxically, the private and public/government employees exerted two times and 2.74 times more likely to be knowledgeable, respectively, than the students in Bangladesh. Despite having much and higher knowledge levels, these two categories of professionals (private and public/government employees) significantly failed to perform proper practice during the second wave of the COVID-19 outbreak in Bangladesh.Similarly, Ferdous et al. [21] demonstrated that the government employees exhibited more accurate knowledge than the students (51.6% vs. 49%); in contrast, private job holders showed less accurate knowledge than the students (47.6% vs. 49%). In another study, Banik et al. [23] found that students showed better practice scores than the govt./private job holder (mean ± standard deviation = 2.52 ± 0.36 vs. 2.47 ± 0.35; p<0.05). Besides, the student category of participants showed around more than two times better practice towards the anti-COVID-19 measures in Bangladesh compared to the businessman (OR = 2.35, 95% CI = 1.01 to 5.67). As the working people/job holders have to go to their offices and outside of the home, they have been more concerned and given instructions and cautionary regarding the COVID-19 from their institutions. On the other hand, students might have a greater opportunity to access the information, but they might be less concerned about COVID-19 by spending time buzzing with friends and others.

Overall, this study seems to corroborate well with other studies conducted in many parts of the world regarding the knowledge, attitude, and practices related to COVID-19. However, it reports that the percentage of Bangladeshi internet users with good knowledge about COVID-19 is lower than the percentage of people in other parts of the world. However, in terms of practices, the results are on par with other countries. The main strength of the study is that it was conducted at a comparatively mature stage of the pandemic as opposed to the initial stage; therefore, it represents the status of the public when more knowledge has been uncovered and some awareness programs have been conducted. So, this study strongly indicates that policies and awareness programs should focus on enhancing the knowledge about COVID-19 among Bangladeshi internet users, particularly the male users.

### Practical implementations

Several anti-epidemic measures such as lockdown, social distancing, isolation, quarantine, and so on are being imposed by national and international authorities to tackle the rapid transmission of SARS-CoV-2 infection. However, the understanding and profound comprehension of the information, severity, and adopted measures are the preconditions for implementing these public health measures. Besides, before assuring the mass vaccination, all the health-related practices against COVID-19 should be established among the common people as a prime concern. The present study revealed a need for more comprehensive educational programs that emphasize uniformity in information from the government and other authorities. A proactive approach to COVID-19 education should be taken, focusing on eliminating misconceptions such as competing viewpoints, myths, and inaccurate information. The results of our study will help public health experts, epidemiologists, and especially those who work on health communication and making an effort to understand the COVID-19 knowledge, attitudes, and practices among the general people to develop cost-effective public health campaigns and education initiatives. The findings obtained in this study might also help the health experts and policy makersdevelop the most successful public health strategy and policy against COVID-19 in the community level of developing countries, including Bangladesh.

### Limitations

This cross-sectional study is not flawless while having some drawbacks which need to be noted. Firstly, the survey was a self-reported web-based study which had no casual inferences. Secondly, the collection of data was not random, rather followed a snowball technique, and therefore this data may not be fully representative of the total population of active internet users in Bangladesh. Thirdly, the sample size is moderate, and higher samples would have provided further strength to the study.

## Conclusion

The proper level of KAP among the general population is mandatorily required for tackling COVID-19 and implementing countrywide anti-COVID-19 measures. The current research’s exact aim was to assess the KAP score among the active and regular internet users during the second wave of the COVID-19 outbreak in Bangladesh. As seen by the current data and evidence, there are significant gaps in awareness, attitudes, and practices among the Bangladeshi population, particularly the males and elders. In addition, the overall percentage of COVID-19 knowledge amongst Bangladeshi internet users is comparatively low compared to other parts of the world. Hence, it is imperative that public health officials take into account these gaps of understanding and try to educate the public as a method in mitigating the effects of the COVID-19 pandemic.

## Data Availability

All the data involved in the study are available in the article. Further raw data will be available from the corresponding author upon reasonable request.

## Acknowledgments

Authors are grateful to the study participants, and the volunteers who supported piloting the surveytool.

## References

[1] N. Chen, M. Zhou, X. Dong, J. Qu, F. Gong, Y. Han, Y. Qiu, J. Wang, Y. Liu, Y. Wei, J. Xia, T. Yu, X. Zhang, L. Zhang, Epidemiological and clinical characteristics of 99 cases of 2019 novel coronavirus pneumonia in Wuhan, China: a descriptive study, The Lancet. 395 (2020) 507–513. https://doi.org/10.1016/S0140-6736(20)30211-7.

[2] Hossain MJ, Kuddus MR, Rashid MA, Sultan MZ. Understanding and dealing the SARS-CoV-2 infection: an updated concise review. Bangladesh Pharm J. 2021;24:61–75.

[3] Weekly epidemiological update. https://covid19.who.int (accessed April 26, 2023).

[4] Hossain MJ, Rahman SMA. Repurposing therapeutic agents against SARS-CoV-2 infection: most promising and neoteric progress. Expert Rev Anti Infect Ther. 2020 Dec 23:1–19. doi: 10.1080/14787210.2021.1864327.

[5] Islam MT, Talukder AK, Siddiqui MN, Islam T. Tackling the COVID-19 pandemic: The Bangladesh perspective, J Public Health Res. 9 (2020). https://doi.org/10.4081/jphr.2020.1794.

[6] Hossain MJ. Social Organizations and Mass Media in COVID-19 Battle: A Bidirectional Approach in Bangladesh. Asia Pac J Public Health. 2021 May;33(4):467–468. doi: 10.1177/10105395211002601.

[7] Coronavirus outbreak: Govt orders closure of public, private offices from March 26 to April 4 | The Daily Star, (n.d.). https://www.thedailystar.net/coronavirus-deadly-new-threat/news/govt-offices-closed-march-26-april-4-cabinet-secretary-1884730 (accessed October 31, 2020).

[8] Islam MR, Jannath S, Moona AA, Akter S, Hossain MJ, Islam SMA. Association between the use of social networking sites and mental health of young generation in Bangladesh: A cross-sectional study. J Community Psychol. 2021 Jul 21. doi: 10.1002/jcop.22675.

[9] World Health Organization (WHO). Accessed July 27, 2021. Available from: Bangladesh: WHO Coronavirus Disease (COVID-19) Dashboard With Vaccination Data | WHO Coronavirus (COVID-19) Dashboard With Vaccination Data

[10] M.K. Al-Hanawi, K. Angawi, N. Alshareef, A.M.N. Qattan, H.Z. Helmy, Y. Abudawood, M. Alqurashi, W.M. Kattan, N.A. Kadasah, G.C. Chirwa, O. Alsharqi, Knowledge, Attitude and Practice Toward COVID-19 Among the Public in the Kingdom of Saudi Arabia: A Cross-Sectional Study. Frontiers in Public Health. 8 (2020). https://doi.org/10.3389/fpubh.2020.00217.

[11] Zhong BL, Luo W, Li HM, Zhang QQ, Liu XG, Li WT, Li Y. Knowledge, attitudes, and practices towards COVID-19 among Chinese residents during the rapid rise period of the COVID-19 outbreak: a quick online cross-sectional survey. Int J Biol Sci. 2020 Mar 15;16(10):1745–1752. doi: 10.7150/ijbs.45221.

[12] E. Hager, I.A. Odetokun, O. Bolarinwa, A. Zainab, O. Okechukwu, A.I. Al-Mustapha, Knowledge, attitude, and perceptions towards the 2019 Coronavirus Pandemic: A bi-national survey in Africa, PLOS ONE. 15 (2020) e0236918. https://doi.org/10.1371/journal.pone.0236918.

[13] D. Souli, M. Dilucca, Knowledge, attitude and practice of secondary school students toward COVID-19 epidemic in Italy: a cross selectional study, BioRxiv. (2020) 2020.05.08.084236. https://doi.org/10.1101/2020.05.08.084236.

[14] Lake EA, Demissie BW, Gebeyehu NA, Wassie AY, Gelaw KA, Azeze GA. Knowledge, attitude and practice towards COVID-19 among health professionals in Ethiopia: A systematic review and meta-analysis. PLoS One. 2021 Feb 19;16(2):e0247204. doi: 10.1371/journal.pone.0247204.

[15] Mistry SK, Ali AM, Yadav UN et al. COVID-19 related misconceptions among older adults in Bangladesh: findings from a cross-sectional study [version 1; peer review: 1 not approved]. F1000Research 2021, 10:216 doi: https://doi.org/10.12688/f1000research.51597.1

[16] Paul A, Sikdar D, Hossain MM, Amin MR, Deeba F, Mahanta J, Jabed MA, Islam MM, Noon SJ, Nath TK. Knowledge, attitudes, and practices toward the novel coronavirus among Bangladeshis: Implications for mitigation measures. PLoS One. 2020 Sep 2;15(9):e0238492. doi: 10.1371/journal.pone.0238492.

[17] Hossain MJ, Kuddus MR, Rahman SMA. Knowledge, Attitudes, and Behavioral Responses Toward COVID-19 During Early Phase in Bangladesh: A Questionnaire-Based Study. Asia Pac J Public Health. 2021 Jan;33(1):141–144. doi: 10.1177/1010539520977328.

[18] Rahman, Shahrul, A. Akter, KF Mostari, S. Ferdousi, IJ Ummon, SM Naafi, Mm Rahman, M. Uddin, S. Tasmin, S. Lopa, SM Sayadat Amin, M. Miah, TK Saha, M. A. Rahim and S. Hossain. “Assessment of knowledge, attitudes and practices towards prevention of coronavirus disease (COVID-19) among Bangladeshi population.” Bangladesh Medical Research Council Bulletin 46 (2020): 73–82.

[19] Hossain MA, Jahid MIK, Hossain KMA, Walton LM, Uddin Z, Haque MO, Kabir MF, Arafat SMY, Sakel M, Faruqui R, Hossain Z. Knowledge, attitudes, and fear of COVID-19 during the Rapid Rise Period in Bangladesh. PLoS One. 2020 Sep 24;15(9):e0239646. doi: 10.1371/journal.pone.0239646.

[20] Rahman, A. and N. J. Sathi. “Knowledge, attitude, and preventive practices toward COVID-19 among Bangladeshi internet users.” (2020).

[21] Ferdous MZ, Islam MS, Sikder MT, Mosaddek ASM, Zegarra-Valdivia JA, Gozal D. Knowledge, attitude, and practice regarding COVID-19 outbreak in Bangladesh: An online-based cross-sectional study. PLoS One. 2020 Oct 9;15(10):e0239254. doi: 10.1371/journal.pone.0239254.

[22] Hossain MB, Alam MZ, Islam MS, Sultan S, Faysal MM, Rima S, Hossain MA, Mahmood MM, Kashfi SS, Mamun AA, Monia HT, Shoma SS. Do knowledge and attitudes matter for preventive behavioral practices toward the COVID-19? A cross-sectional online survey among the adult population in Bangladesh. Heliyon. 2020 Dec;6(12):e05799. doi: 10.1016/j.heliyon.2020.e05799.

[23] Banik R, Rahman M, Sikder MT, Rahman QM, Pranta MUR. Knowledge, attitudes, and practices related to the COVID-19 pandemic among Bangladeshi youth: a web-based cross-sectional analysis. Z Gesundh Wiss. 2021 Jan 16:1–11. doi: 10.1007/s10389-020-01432-7.

[24] Islam S, Emran GI, Rahman E, Banik R, Sikder T, Smith L, Hossain S. Knowledge, attitudes and practices associated with the COVID-19 among slum dwellers resided in Dhaka City: a Bangladeshi interview-based survey. J Public Health (Oxf). 2021 Apr 12;43(1):13–25. doi: 10.1093/pubmed/fdaa182.

[25] Maude RR, Jongdeepaisal M, Skuntaniyom S, Muntajit T, Blacksell SD, Khuenpetch W, Pan-Ngum W, Taleangkaphan K, Malathum K, Maude RJ. Improving knowledge, attitudes and practice to prevent COVID-19 transmission in healthcare workers and the public in Thailand. BMC Public Health. 2021 Apr 18;21(1):749. doi: 10.1186/s12889-021-10768-y.

[26] Hasan, Md Zahid, AM Rumayan Hasan, Md Golam Rabbani, Mohammad Abdus Selim, and Shehrin Shaila Mahmood. “Knowledge, attitude, and practice of Bangladeshi urban slum dwellers towards COVID-19 transmission-prevention: A cross-sectional study.” PLOS Global Public Health 2, no. 9 (2022): e0001017.

[27] R.C. Reuben, M.M.A. Danladi, D.A. Saleh, P.E. Ejembi, Knowledge, Attitudes and Practices Towards COVID-19: An Epidemiological Survey in North-Central Nigeria, J Community Health. (2020) 1–14. https://doi.org/10.1007/s10900-020-00881-1.

[28] L.A. Goodman, Snowball Sampling, Ann. Math. Statist. 32 (1961) 148–170. https://doi.org/10.1214/aoms/1177705148.

[29] Hossain, M. J., Ahmmed, F., Rahman, S. M. A., Sanam, S., Emran, T. B., Mitra, S. Impact of online education on fear of academic delay and psychological distress among university students following one year of the COVID-19 outbreak in Bangladesh. Heliyon 2021. doi: https://doi.org/10.1016/j.heliyon.2021.e07388.

[30] Andrade C, Menon V, Ameen S, Kumar Praharaj S. Designing and Conducting Knowledge, Attitude, and Practice Surveys in Psychiatry: Practical Guidance. Indian J Psychol Med. 2020 Aug 27;42(5):478–481. doi: 10.1177/0253717620946111.

[31] Kaliyaperumal, K. Guideline for conducting a knowledge, attitude and practice (KAP) study. Community Ophthalmology 2004; 4: 7–9

[32] Hossain MJ, Hridoy A, Rahman SMA, Ahmmed F. Major Depressive and Generalized Anxiety Disorders Among University Students During the Second Wave of COVID-19 Outbreak in Bangladesh. Asia Pac J Public Health. 2021 May 10:10105395211014345. doi: 10.1177/10105395211014345. Epub ahead of print. PMID: 33969713.

[33] World Medical Association Declaration of Helsinki. Ethical principles for medical research involving human subjects., Bull World Health Organ. 79 (2001) 373–374.

[34] Hossain MJ. Is Bangladesh moving toward herd immunity? Current COVID-19 perspective. Bangladesh J Infect Dis. 2020;7(suppl_2): S63–S66.

[35] For the first time, India has more rural net users than urban - The Times Of India - Bangalore, 5/6/2020, (n.d.). https://epaper.timesgroup.com/Olive/ODN/TimesOfIndia/shared/ShowArticle.aspx?doc=TOIBG%2F2020%2F05%2F06&entity=Ar00110&sk=0E13F922&mode=text# (accessed October 31, 2020).

[36] Sharma M, Yadav K, Yadav N, Ferdinand KC. Zika virus pandemic-analysis of Facebook as a social media health information platform. Am J Infect Control. 2017 Mar 1;45(3):301–302. doi: 10.1016/j.ajic.2016.08.022.

[37] Rabbani, Md Golam, Orin Akter, Md Zahid Hasan, Nandeeta Samad, Shehrin Shaila Mahmood, and Taufique Joarder. “COVID-19 Knowledge, Attitudes, and Practices Among People in Bangladesh: Telephone-Based Cross-sectional Survey.” JMIR Formative Research 5, no. 11 (2021): e28344.

[38] Islam, Md, Faroque Ahmed, and Afrin Sadia Rumana. “Creative social media use for Covid-19 prevention in Bangladesh: a structural equation modeling approach.” Social Network Analysis and Mining 11, no. 1 (2021): 1–14.

[39] Hayat K, Rosenthal M, Xu S, Arshed M, Li P, Zhai P, Desalegn GK, Fang Y. View of Pakistani Residents toward Coronavirus Disease (COVID-19) during a Rapid Outbreak: A Rapid Online Survey. Int J Environ Res Public Health. 2020 May 12;17(10):3347. doi: 10.3390/ijerph17103347.

[40] Roy D, Tripathy S, Kar SK, Sharma N, Verma SK, Kaushal V. Study of knowledge, attitude, anxiety & perceived mental healthcare need in Indian population during COVID-19 pandemic. Asian J Psychiatr. 2020 Jun;51:102083. doi: 10.1016/j.ajp.2020.102083.

[41] Azlan AA, Hamzah MR, Sern TJ, Ayub SH, Mohamad E. Public knowledge, attitudes and practices towards COVID-19: A cross-sectional study in Malaysia. PLoS One. 2020 May 21;15(5):e0233668. doi: 10.1371/journal.pone.0233668.

[42] Honarvar B, Lankarani KB, Kharmandar A, Shaygani F, Zahedroozgar M, Rahmanian Haghighi MR, Ghahramani S, Honarvar H, Daryabadi MM, Salavati Z, Hashemi SM, Joulaei H, Zare M. Knowledge, attitudes, risk perceptions, and practices of adults toward COVID-19: a population and field-based study from Iran. Int J Public Health. 2020 Jul;65(6):731–739. doi: 10.1007/s00038-020-01406-2.

[43] Hezima A, Aljafari A, Aljafari A, Mohammad A, Adel I. Knowledge, attitudes, and practices of Sudanese residents towards COVID-19. East Mediterr Health J. 2020 Jun 24;26(6):646–651. doi: 10.26719/emhj.20.076.

[44] Ck E, Ai W, Va K. Knowledge, Attitude and Practice of Clients towards COVID-19 at Primary Healthcare Facilities in Rivers State, Nigeria, (2020). https://doi.org/10.21203/rs.3.rs-40966/v1.

[45] Asmelash D, Fasil A, Tegegne Y, Akalu TY, Ferede HA, Aynalem GL. Knowledge, Attitudes and Practices Toward Prevention and Early Detection of COVID-19 and Associated Factors Among Religious Clerics and Traditional Healers in Gondar Town, Northwest Ethiopia: A Community-Based Study. Risk Manag Healthc Policy. 2020 Oct 20;13:2239–2250. doi: 10.2147/RMHP.S277846.

[46] Chen X, Chen H. Differences in Preventive Behaviors of COVID-19 between Urban and Rural Residents: Lessons Learned from A Cross-Sectional Study in China. Int J Environ Res Public Health. 2020 Jun 20;17(12):4437. doi: 10.3390/ijerph17124437.

[47] Wadood MA, Mamun ASMA, Rafi MA, Islam MK, Mohd S, Lee LL, Hossain MG. Knowledge, attitude, practice and perception regarding COVID-19 among students in Bangladesh: Survey in Rajshahi University. medRxiv. 2020. doi: https://doi.org/10.1101/2020.04.21.20074757

[48] Abdelhafiz AS, Mohammed Z, Ibrahim ME, Ziady HH, Alorabi M, Ayyad M, Sultan EA. Knowledge, Perceptions, and Attitude of Egyptians Towards the Novel Coronavirus Disease (COVID-19). J Community Health. 2020 Oct;45(5):881–890. doi: 10.1007/s10900-020-00827-7.

[49] Aynalem YA, Akalu TY, Gebresellassie B, Sharew NT, Shiferaw WS. Assessment of undergraduate student knowledge, practices, and attitude towards COVID-19 in Debre Berhan University, Ethiopia. Research Square 2020. DOI: 10.21203/rs.3.rs-28556/v1.

